# *De novo* and inherited variants in *DDX39B* cause a Novel Syndrome Characterized by Neurodevelopmental Delay, Short Stature, and Congenital Hypotonia

**DOI:** 10.1101/2023.07.15.23292630

**Authors:** Kevin T.A. Booth, Sharayu V. Jangam, Martin M.C. Chui, Kayla Treat, Lorenzo Graziani, Alessia Soldano, Kerry White, Celanie K. Christensen, Ty Lynnes, Shinya Yamamoto, Oguz Kanca, Mandy H.Y. Tsang, Sally A. Lynch, Sureni V. Mullegama, Julia Batista, Daniela Iancu, Shelag K. Joss, Christopher C.Y. Mak, Anna Y. Kwong, Hugo J. Bellen, BCM Center for Precision Medicine Models, Erin Conboy, Remo Sanges, Michael F. Wangler, Brian H.Y. Chung, Francesco Vetrini

**Affiliations:** Indiana University School of Medicine; Department of Medical and Molecular Genetics; Undiagnosed Rare Disease Clinic, Indianapolis, Indiana, USA; Indiana University School of Medicine, Indianapolis, Indiana, USA; Department of Otolaryngology-Head and Neck Surgery, Indiana University School of Medicine, Indianapolis, Indiana, USA; Riley Developmental Pediatrics, Indianapolis, IN, USA; Department of Molecular and Human Genetics, Baylor College of Medicine; Houston, TX, USA; Center for Precision Medicine Modeling, Baylor College of Medicine; Houston, TX, USA; Jan and Dan Duncan Neurological Research Institute, Texas Children’s Hospital, Houston, TX, USA; Department of Paediatrics and Adolescent Medicine, School of Clinical Medicine, LKS Faculty of Medicine, The University of Hong Kong, HK; University of Plymouth, UK; Children’s Health Ireland, Crumlin, Dublin, Republic of Ireland; University College of London, U.K.; Queen Elizabeth University Hospital, Glasgow, U.K.; GeneDx, Gaithersburg, MD, USA; Scuola Internazionale Superiore di Studi Avanzati, Trieste, Italy

## Abstract

*DDX39B* is a member of the DEAD-box family of ATP-dependent RNA helicases. DEAD-box proteins are ubiquitously expressed from yeast to humans and perform essential functions associated with mRNA metabolism. *DDX39B* is also a crucial component of the TRanscription-EXport (TREX) super protein complex, which recent studies have highlighted the important role of its subunits in neurodevelopmental disorders. Here, we describe six individuals from five families, four with novel *de novo* missense variants in *DDX39B,* and one carrying an inherited splicing variant, all presenting with mild to severe global developmental delay, congenital hypotonia, epilepsy, short stature, skeletal abnormalities and variable dysmorphic features. 3D molecular modeling predicts these variants would alter protein structure. *DDX39B* is a conserved gene and *Drosophila melanogaster* (fruit flies) studies were conducted. We generated a new Hel25E Kozak-GAL4 allele which disrupts the fly gene and allows expression of transgenes. We also generated transgenic *DDX39B*-reference and variant flies. However, human reference *DDX39B* when overexpressed ubiquitously leads to lethality but the variants found in the patients do not recapitulate the lethality suggesting that the mutants are loss of function alleles. Blood transcriptomics revealed a significant excess of aberrant splicing events, indicating a disrupted mRNA processing as anticipated from the role of *DDX39B* in mRNA metabolism. Our human genetic data, coupled with *in silico* and *in vivo* data supports that *DDX39B* is a novel candidate gene in a potential group of disorders named TREX-complex-related neurodevelopmental syndrome.

## Introduction

Pre-mRNA splicing and mRNA export from the nucleus, are highly coordinated and tightly regulated processes. Both processes require protein complexes’ assembly, coordination, and synergy for proper function. Defects in the proteins required for either or both processes can result in abnormal mRNA processing and, ultimately, disease.

The *DDX39B* gene (MIM: 142560, RefSeq NM_004640.7) encodes for the 428 amino acid DExD/H-box family ATP-dependent RNA helicase DDX39B protein. DDX39B is ubiquitously present in organisms from yeast to human, and is an essential splicing factor with a variety of functions in the pre-spliceosome and mature spliceosome assembly.^1^ DDX39B is also a key member of the TRanscription-EXport (TREX) super protein complex, where it mediates the THO-complex assembly, ATP hydrolysis, and recruiting export adapter ALYREF to THO-DDX39B bound pre-mRNA.^2^ As part of the TREX, DDX39B plays a diverse role in mRNA physiology, including pre-mRNA splicing, nuclear export, ribosome assembly, and translation initiation.^3–6^ Pathogenic variants in proteins comprising the TREX complex have been causally linked to neurodevelopment and neurodegenerative diseases^7–9^, such as *THOC2* (MIM: 300395, RefSeq NM_001081550.2) which is associated with the X-linked intellectual developmental disorder 12 (MIM: 300957) and *THOC6* (MIM: 615403, RefSeq NM_024339.5) that is responsible for Beaulieu-Boycott-Innes syndrome (MIM: 613680).

Here we implicate *DDX39B* in a novel neurodevelopmental disorder. We describe a cohort of six individuals from five families affected by mild to severe global developmental delay, hypotonia, history of epilepsy or seizure, short stature, skeletal abnormalities, and variable dysmorphic features, with disease-causing *de novo* missense variants or inherited splice-altering variants in *DDX39B*. We also employ *Drosophila* models and RNA sequencing on the patient’s blood sample to study the variants and provide evidence for their role in the neurodevelopmental phenotypes observed in the clinic.

## Materials and methods

### Subjects

All The procedures were followed in accordance with the ethical standards of the respective institutions. Proper informed consent was obtained from legal guardians of affected individuals. The probands’ parents or legal guardians consented to the publication of their identifiable photos and detailed clinical presentation. **[*As per medRxiv policy, the whole and detailed case history for the six patients, as well as the patients’ images, have been removed. To obtain more detailed information, please contact the authors]*.**

### DNA Sequencing

The exome sequencing (ES) analyses were conducted on a clinical or research basis. For proband A, genomic DNA from the proband and parents was used, the exonic regions and flanking splice junctions of the genome were captured using the IDT xGen Exome Research Panel v1.0 (Integrated DNA Technologies, Coralville, IA). Massively parallel (NextGen) sequencing was done on an Illumina system with 100bp or greater paired end reads. Reads were aligned to human genome build GRCh37/UCSC hg19, and analyzed for sequence variants using a custom-developed analysis tool. The general assertion criteria for variant classification are publicly available on the GeneDx ClinVar submission page (http://www.ncbi.nlm.nih.gov/clinvar/submitters/26957/)^10^ For proband B: Trio ES was performed using the Twist Human Core Exome Kit (Twist Bioscience, San Francisco, CA, USA). Sequencing of paired end 150bp reads was performed on a NextSeq500 (Illumina, SanDiego, CA, USA). Subsequent bioinformatic analysis: read mapping, variant calling and quality filtering, variant filtering and prioritization was performed as described^11^. For Proband C, genomic DNA from proband and parents were obtained from the peripheral blood using Qiagen Blood Mini Kit. Trio exome libraries were prepared using Illumina TruSeq exome for proband and father, and IDT xGen xome for mother. Sequencing was performed using Illumina NextSeq 500 platform. Sequence reads were aligned to human genome build hg19. In-house bioinformatics pipeline was used for variant calling and analysis. The identification of Proband D and Family 1 are a result of a Matchmaker exchange query (DECIPHER: 264142 and Family1: https://www.deciphergenomics.org/ddd/research-variant/f464bfb477f6d25863d934c3f9593ece/overview). Sequence generation, variant identification and prioritization for Proband D and Family 1 was performed as described [INSERT DDD REF].

### 3D modeling

To explore the impact of the missense variants identified in this study we employed 3D protein modeling as described^12^ Briefly, a Alphafold2 model of the DDX39B protein (AF-Q13838-F1) containing residues 1-428 was obtain. Pymol was used to visualize the wildtype model, perform mutagenesis, identify H-bonds and take measurements.

### Transcriptomics

Peripheral blood of Proband C was treated with red blood cell lysis buffer (Qiagen) and white blood cells were collected for blood RNA extraction using TRIzol™ reagent (Thermo Fisher Scientific) according to manufacturers’ instruction. The quality of the extracted RNA was confirmed with an RNA integrity number (RIN) ≥ 8. Non-strand-specific cDNA library was prepared from the sample using a poly-A-tail capturing method with the KAPA mRNA HyperPrep Kit (Roche Diagnostics) according to manufacturer’s protocol, and subsequently sequenced as 150-bp paired-end runs on an Illumina NovaSeq 6000 platform with at least 100 million reads per sample at the Center for PanorOmic Sciences (CPOS), University of Hong Kong. Data cleaning of the transcriptomics profile in Fastq format was performed by AfterQC (v0.9.6) while the sequencing data was mapped to the hg19/GRCh37 human reference genome and outputted as bam files using STAR (v2.5.2a) in two-pass mode. The sequencing data were processed with DROP v1.1.0, which integrates statistical modules to detect significant aberrant expression, aberrant splicing and monoallelic expression.^13^ In particular, aberrant splicing events were detected using the FRASER module in DROP, an annotation-free algorithm for the detection of novel splice sites4. In FRASER, split (representing an exon–exon junction) and non-split (spanning the splice sites and retaining introns) reads were counted for each transcripts. Statistical metrics indicating the splicing events was calculated based on the corresponding ratio between the split and non-split reads, and quantified into parameters of alternative acceptors (ψ5), alternative donors (ψ3), and splicing efficiency (θ). Aberrant splicing events were defined as false discovery rate ≤ 0.05 and |Δψ| or |Δθ| ≥ 0.2, and were further evaluated as mentioned in the Result and Discussion sections.

### Drosophila Experiments

#### Fly husbandry

Stocks were raised on standard fly food (water, yeast, soy flour, cornmeal, agar, molasses, and propionic acid) at room temperature (~22°C) and routinely maintained. Unless otherwise stated, all flies used in experiments were grown in a temperature and humidity-controlled incubator at 25°C and 50% humidity on a 12-hour light/dark cycle. The y[1] w[67c23]; P[w[+mC]=lacW]Hel25E[k11511]/CyO and the w[1118]; Df(2L)BSC167/CyO fly stocks were ascertained from the Bloomington Stock Center stock numbers #11053 and #9602, respectively. The p[Actin-GAL4] balanced over CyO-Tb stock was generated as described.^14^ The stocks p[GawB]elav[C155], p[daughterless-GAL4] and p[eyelessGAL4] were generous gifts from Hugo Bellen’s Lab.

#### Generation of the DDX39B transgenic flies under UAS promoter

Reference human cDNA “IOH3681” from the Ken Scott collection was used for these constructs. By designing the site-specific primers, each variant was generated with a Q5 mutagenesis protocol (NEB-E0554S). After confirming via Sanger sequencing analysis, the constructs in the pENTR-221 vector were cloned into pGW-[UASg] destination vector by using LR Clonase™ II (Thermo Fischer-11791020). Sequence-verified pattB[UAS-DDX39Bref], pattB[UAS-DDX39BG37C], pattB[UAS-DDX39BG92D] & pattB[UAS-DDx39BR123Q] constructs were microinjected into VK00033 docking site embryos to generate pBac[UAS-DDX39Bref/Var]VK00033 lines.

### Humanization assay

#### Over-expression assay (Assessment of lethality and morphological phenotypes)

Lethality and morphological phenotyping assays were performed by crossing GAL4 drivers as indicated in the text, using 5-10 virgin females crossed to a similar number of males. After every 3-5 days parents were transferred into a new vial to collect multiple F1-progenies. Flies were collected after most pupae enclosed, and the total number of flies was scored based on the presence or absence of balancers. For the lethality assessment, a minimum of 100 flies were scored. Viability was calculated via evaluation of the number of observed progenies compared to the number of expected progeny based on the mendelian ratio. Animals were classified as lethal if the O/E ratio was less than 0.15, and semi-lethality is classified as an O/E ratio less than 0.8. Assessment of morphological phenotypes was only done for animals lacking balancers, and phenotypes were noted if they appeared in more than 70% of the progeny.

## Results

### Cohort and Variant Identification

This international collaboration was facilitated by GeneMatcher (GeneMatcher, https://genematcher.org/) and the variants were identified by trio-based ES in diagnostic or research setting. Written informed consent was obtained from the parents or legal guardians of all study participants. The six individuals reported here range in age from 3 years to 36 years, and half of them are adults (>18 years). These six patients present with mild to severe motor developmental delay and intellectual disabilities, with hypotonia observed in all individuals (Table 1). Additional neurological phenotypes include history of epilepsy or seizure (n=4/6) and autism behaviors (n=3/6). Abnormalities in growth parameters is a common shared feature, including short stature (< 3rd centile, n=4/6) and head circumference (n=3/6). Dysmorphic features observed among the cohort are ptosis (n=2/6), upslanted palpebral fissures (n=3/6), downturned corners of the mouth (n=2/6), epicanthic folds (n=2/6), and esotropia (n=2/6). Five individuals in the cohort have skeletal abnormalities, especially lateral deviation at the metatarsophalangeal joint (n=4/6). ***[As per medRxiv policy, the whole and detailed case history for the six patients, as well as the patients’ images, have been removed. To obtain more detailed information, please contact the authors]*.**

After informed and written consent, the proband A and her parents underwent trio Clinical ES, revealing a *de novo* c.109G>T p.(Gly37Cys) missense variant in *DDX39B*. No additional variants were identified that could potentially explain the patient’s clinical presentation. Through GeneMatcher, we identified two additional patients with overlapping clinical features harboring *de novo* missense changes in *DDX39B*, *novo* c.257G>A p.(Gly92Asp) and c.368G>A p.(Arg123Gln) variant. Proband D, previously reported in the Deciphering Developmental Disorders Study^15^ harbors a *de novo* c.132C>G p.(Ser44Arg) variant in *DDX39B*. This is the first in-depth look at the patient’s phenotype and the p.(Ser44Arg) variant in *DDX39B*. Finally, Family 1, identified through the Deciphering Developmental Disorders Study, is a mother and a son carrying a variant at the exon 5 splicing acceptor site (c.433-1G>T).

No additional variants, either in known or novel genes, have been identified as potential candidates in the exome analysis of the above cohort. Detailed molecular features of the DDX39B genomic variants are summarized Figure 1.

**Figure 1.**
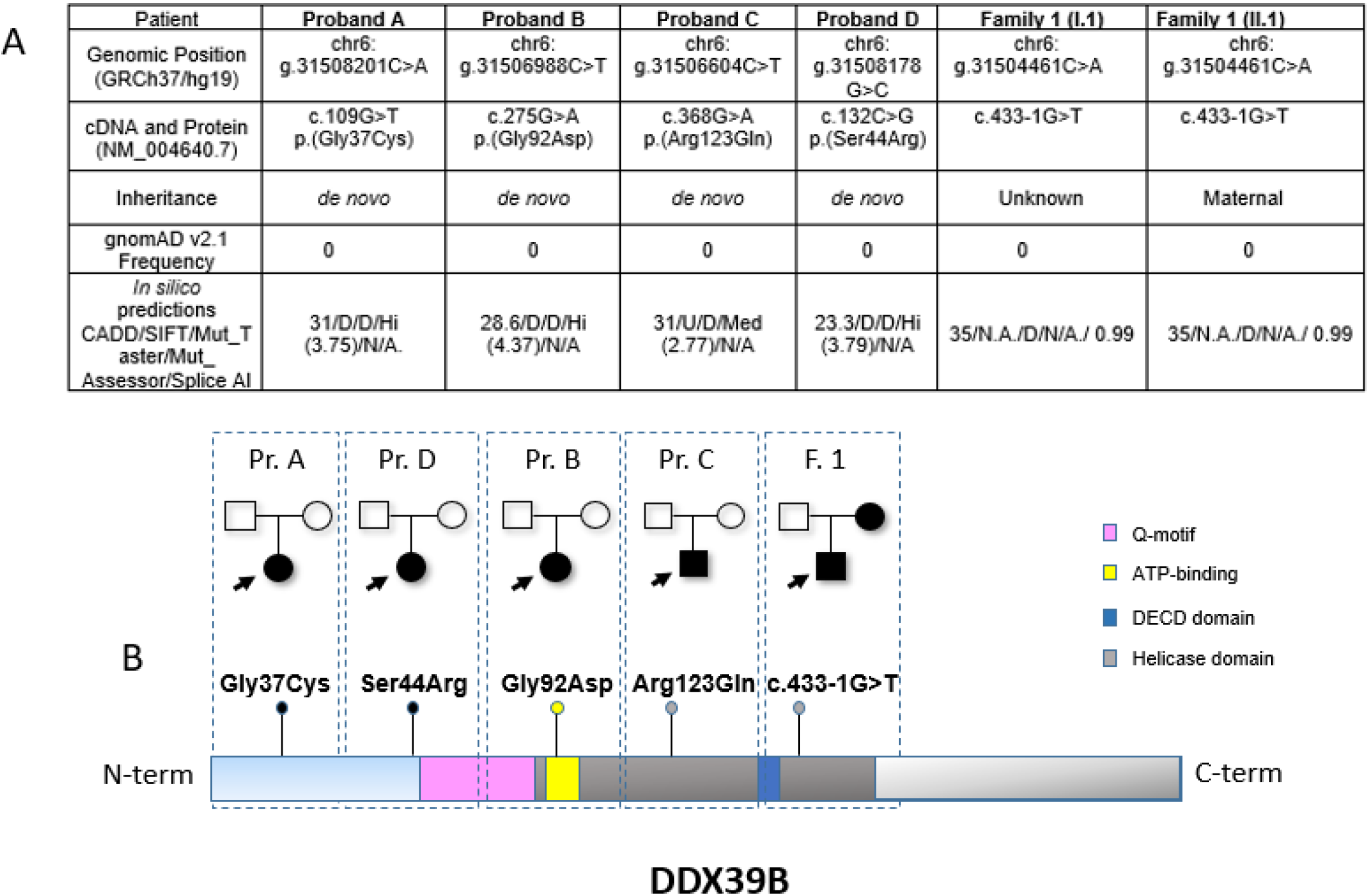
DDX39B molecular findings. (A) Table summarizing the molecular findings in the six patients. (B) Schematic representation of DDX39B protein domains. The locations of all the variants described in this study are annotated above the schematic and the annotation of the functional domains are based on multiple studies.^21,23,55^

### Variant analysis and *in silico* analysis

To assess the pathogenic effect of the *DDX39B* variants, we evaluated the genetic data first and gathered information from multiple sources, including Genome Aggregation Database (gnomAD)^16^, and used multiple *in silico* predicting tools (Figure 1A). All five variants identified are absent from gnomAD v2.1, and four have been confirmed *de novo*. *DDX39B* constraint analyses in gnomAD indicate that the gene is intolerant to loss of function (LoF) variants (pLI=1) and is highly intolerant to missense alterations across the entire gene (Z score = 4.564).^16^ All four missense variants are predicted deleterious by *in silico* prediction tools (Figure 1A). Combined Annotation Dependent Depletion (CADD) scores of the variants in our patients are all above 23.3, where scores above 20 represent variants that are among the 1% most deleterious.^17^

DDX39B is a highly conserved protein from yeast to humans and all four missense variants described here, impact highly conserved residues (Figure 2A and 2B). We evaluated the amino acid location of the four missense variants in gnomAD. All the variants were found in missense-depleted regions, also previously described ‘desert stretches^18^, corresponding to critical domains of the protein. We also performed MetaDome analysis (https://stuart.radboudumc.nl/metadome) which integrates publicly available data of human genetic variation to predict the pathogenicity of genetic variants through aggregation of homologous human protein domains.^19^ MetaDome showed that all the missense changes affecting residues Gly37, Ser44, Gly92 and Arg123 are intolerant or highly intolerant. Moreover, residues Gly92 and Arg123 are highly conserved in both *DDX59* (MIM: 174300) and *DDX6* (MIM: 618653), other DExD/H-box RNA helicase family members implicated in neurodevelopmental disorders and appear to be important residues involved in ATPase activity (<u>G</u>TK triplet, Motif I) and RNA binding ability (invariant domain PT<u>R</u>ELA Motif Ia) (Figure 1B and 2B-D), respectively.^20–23^

**Figure 2.**
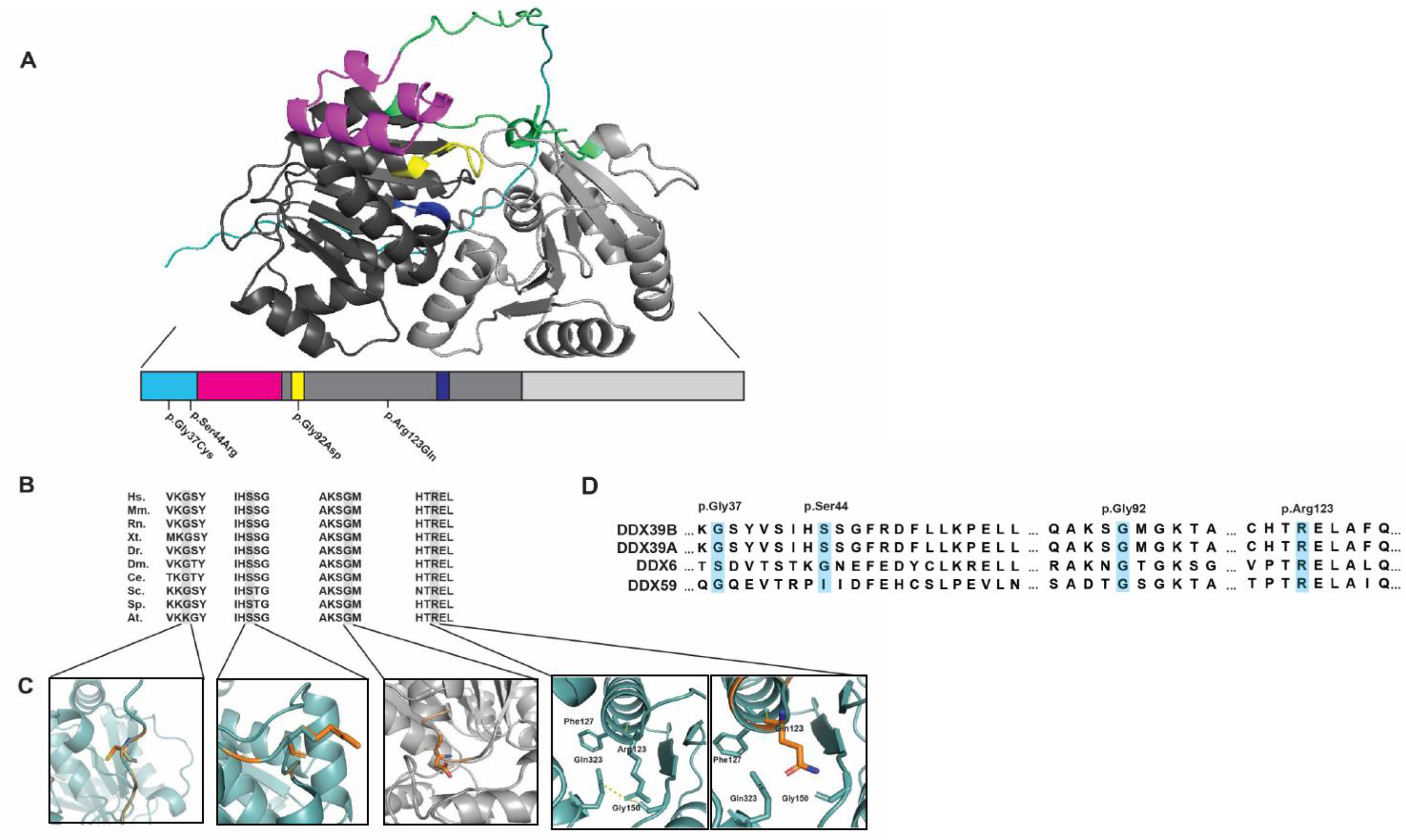
Protein schematic, alignment and 3D modeling of DDX39B. (A) DDX39B is a highly conserved RNA helicase consisting of a disordered region (cyan), Q-motif (pink), Helicase ATP binding domain (light-grey), and a helicase c-terminal domain (dark-grey). Housed inside the Helicase ATP binding domain is the ATP-binding pocket (yellow, residues 89-96) and a DECD domain (blue, residues 196-199). (B) DDX39B conservation across species. (C) 3D protein modeling of the four de novo missense variants identified in this study. In grey is the wildtype, and orange represents the mutant residue. (Abbreviations: Hs., Homo sapiens; Mm, Mus musculus; Rn, Rattus norvegicus; Xt., Xenopus tropicalis; Dr., Danio rerio; Dm. Drosophila melanogaster; Ce., Caenorhabditis elegans; Sc., Saccharomyces cerevisiae; Sp., Schizosaccharomyces pombe; At., Arabidopsis thaliana). (D) DDX39B paralog conservation, highlighted are the residues affected by missense changes.

### 3D protein modeling

To explore the functional impact of the missense variants identified in this study, first, we employed 3D protein modeling. All four missense variants are predicted to impact local structure and folding. The Gly37 is in the N-terminus disordered tail region. Its substitution for a bulkier and more rigid cystine does not alter any hydrogen bonds; however, the ridged cystine significantly alters the local configuration (Figure 2C). The Ser44 is located one residue outside the Q-motif, which is essential for ATP binding and hydrolysis. The p.(Ser44Arg) changes a small uncharged serine with a bulky and positively charged arginine, alters local folding confirmation, and may impair ATP binding or hydrolysis. The wildtype Ser44 residue is predicted to undergo posttranslational modification to phosphoserine.^24^ The p.(Ser44Arg) change is expected to abolish this modification. The Gly92 is in the ATP-binding pocket (residues 89-96). Replacing the small glycine with a large negatively charged aspartic acid alters local conformation and likely impedes DDX39B’s ATP hydrolysis. Finally, substituting the large positively charged Arg123 in the helicase ATP binding domain with a non-polar glutamine results in the loss of two hydrogen bonds with residues Gly150 and Gln323 (Figure 2C). The loss of the hydrogen bonds and the change in charge alters local configuration and is predicted to impact global protein folding.

#### Drosophila in vivo analysis

To investigate the impact of the missense variants in DDX39B *in vivo*, we utilized *Drosophila* as the model organism. We generated a *Hel25E*-specific T2A-GAL4 construct which replaces the entire coding region of the *Hel25E* to KozakT2A-GAL4 (Figure 3A and 3B). This *Hel25E^Kozak-GAL4^*is a loss-of-function allele and leads to GAL4-mediated expression in a similar spatial and temporal expression pattern to determine gene expression or carry out rescue experiments as described.^14^ Breifly, the GAL4 transcriptional activator protein binds to the upstream activation sequence (UAS) containing constructs to drive the expression of a reporter protein.^25^ When a nuclear-localized mCherry reporter line (UAS-mCherry.nls) was crossed with the *Hel25E^Kozak-GAL4^*, the resulting larvae were dissected co-stained with neuronal marker-*ELAV* and glial marker-*repo*. The mCherry.nls signal robustly colocalized with most neurons and glia, suggesting that the *Hel25E* is expressed in neurons and glia during the developing stages (Figure 3B’).

**Figure 3.**
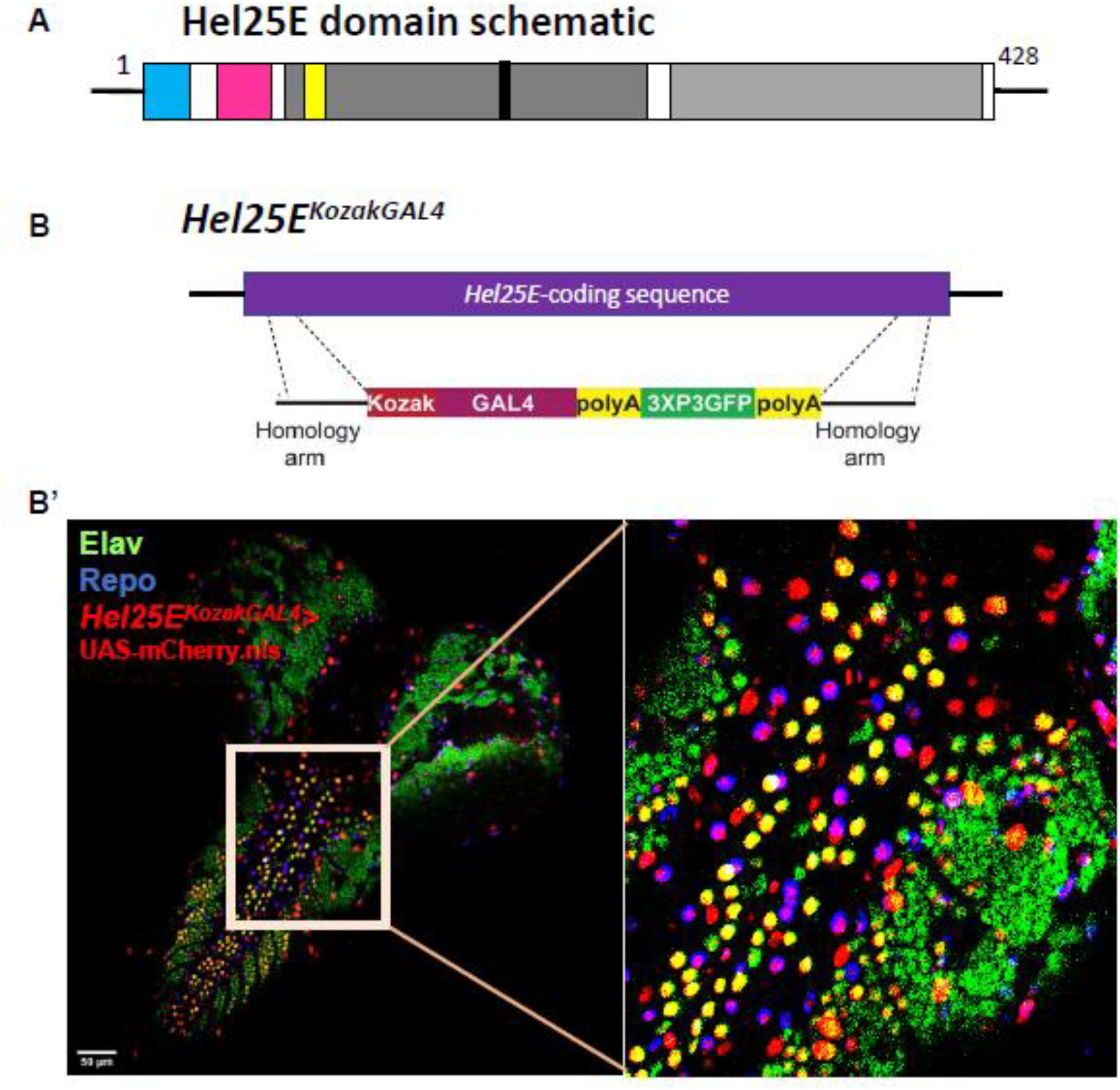
*Drosophila* homolog *Hel25E*. (A) Domain structure schematic of Hel25E protein. The color code matches the DDX39B schematic represented in figure 2. (A’) The *Kozak-GAL4* insertion schematic. (B) Expression study of the *Hel25E^KozakGAL4^*>UAS-mCherry.nls in the third instar larval brain co-stained with Neuronal marker-Elav and Glial marker-Repo. Scale bar=50um. The inset is the zoomed version of the exact same figure.

After confirmation of the constructs, via Sanger sequencing, all four constructs, including the reference, were injected into fly embryos to generate the transgenic flies under the *UAS* promotor as *UAS-DDX39B^ref/Var^* (see Materials and Methods) (Figure 4A). When these *UAS-DDX39B* constructs were crossed with *Hel25E^Kozak-GAL4^*we observed that neither reference nor variant human *DDX39B* cDNA were able to rescue the fly null lethality (Figure 4B). However, when the human cDNAs were overexpressed in the fly wildtype background with ubiquitous *GAL4* lines as *Actin-GAL4 or daughterless-GAL4*, we observed that the *DDx39B*^reference^ was lethal, suggesting toxicity of the human gene in flies. Interestingly, the variants identified in the human subjects did not have this same toxic impact seen in the reference human *DDX39B*. Indeed, overexpression of the UAS-*DDx39B^G37C^*, *DDx39B*^G92D,^ or *DDx39B*^R123Q^, when crossed with the *ubiquitous-GAL4* lines, resulted in phenotypically normal adult flies with standard observed/ expected Mendelian ratio. When tissue-specific GAL4 lines were used, like neuronal-specific *elav-GAL4* and eyes-specific *eyeless-GAL4*, we did not notice any apparent phenotypic change in any DDX39B expressing F1 (Figure 4C). Taken together, these results suggest that the human reference DDX39B is toxic in flies when overexpressed, and the variants remove that toxic effect. These findings support a loss-of-function effect for the human variants.

**Figure 4.**
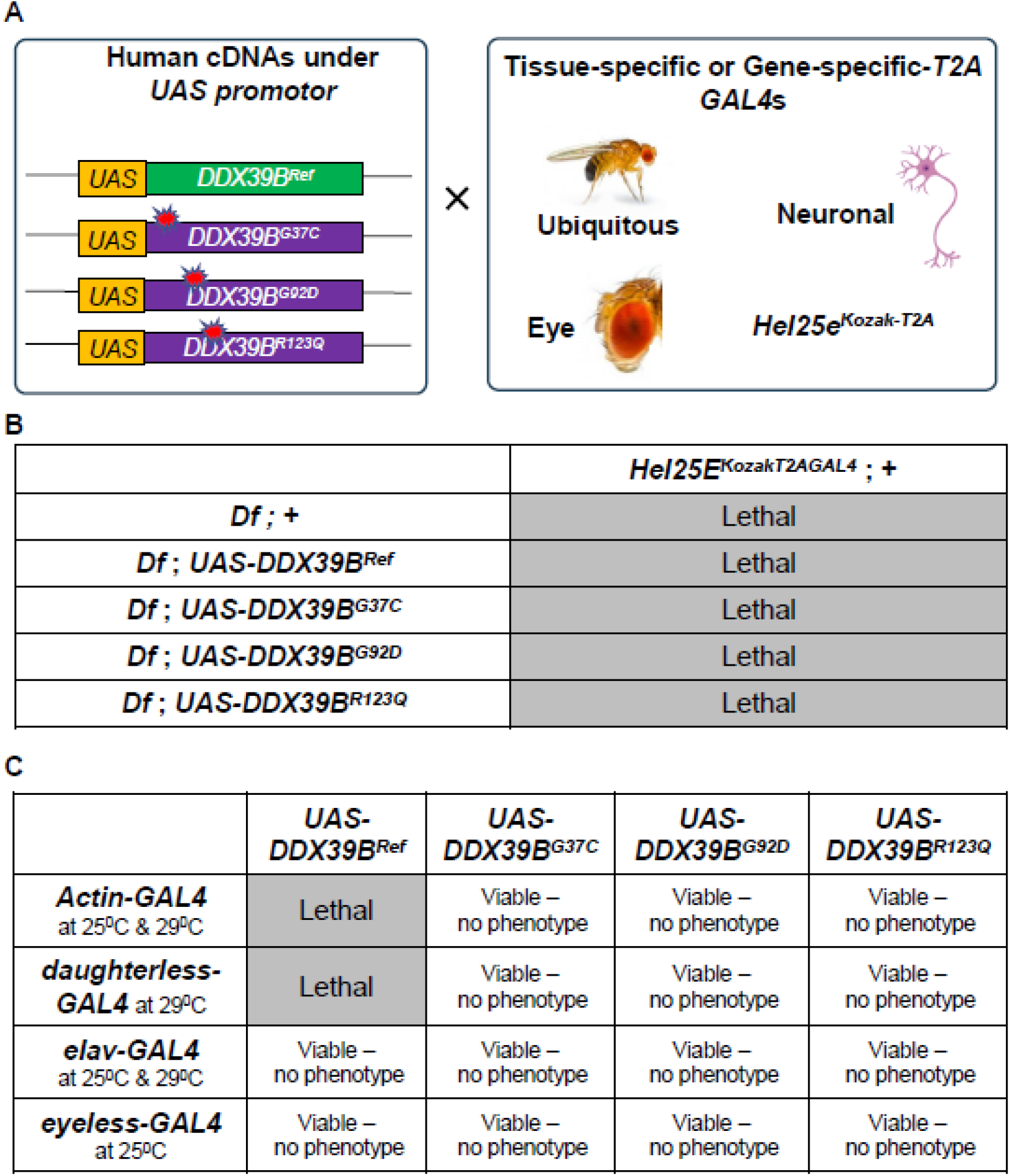
Human cDNA transgenic study in vivo. (A) Schematic representation of the different variant-specific construct-making under the *UAS* promotor. These transgenic flies were crossed with different tissue-specific and gene-specific *GAL4* lines. (B) Humanization/rescue assay-human DDX39B transgenics are expressed in the fly null background. (C) Overexpression assay-human DDX39B transgenics were expressed spacially and temporally in the flies with different *GAL4* lines at different temperatures in the fly wildtype background.

### Global Transcriptomic Profiling

To further support the impact of altered *DDX39B* protein on the global transcriptomics profile in the affected individuals, we performed RNA sequencing (RNA-seq) on the blood sample of Proband C. RNA sample extracted from peripheral blood of the patient was used for preparation of non-strand-specific RNA library with poly-A tailed capture method, according to manufacturer’ protocols. RNA libraries were sequenced as paired-end runs with over 100 million reads per sample. Then sequenced data were mapped to hg19/GRCh37 human reference genome using STAR (v2.5.2a) in two-pass mode, followed by processing and statistical analysis of the “Detection of RNA Outliers Pipeline” (DROP)^13^. As the *DDX39B* variant is present only in the patient, we expect the transcriptomics profile of our patient is unique. Therefore, we employed an “outlier approach” to detect any aberrant events in the transcriptome of the affected individual (n=1) that deviated from our internal RNA-seq samples and external GTEx controls (n=85). We identified 99 aberrant splicing events in the transcriptome of Proband C, with a false discovery rate ≤ 0.05 and magnitude of splicing metrics (psi3 for alternative splicing donors, psi5 for alternative splicing acceptors, and theta for aberrant splicing efficiencies) ≥ 0.2 (Figure 5A-C). The number of aberrant splicing events in Proband C is significantly larger than in our internal samples and external GTEx controls (Z score among internal samples: 3.41, p<0.01 by two-tailed Student’s T-test; Figure 5D), suggesting a generally disrupted transcriptomics profile.

**Figure 5.**
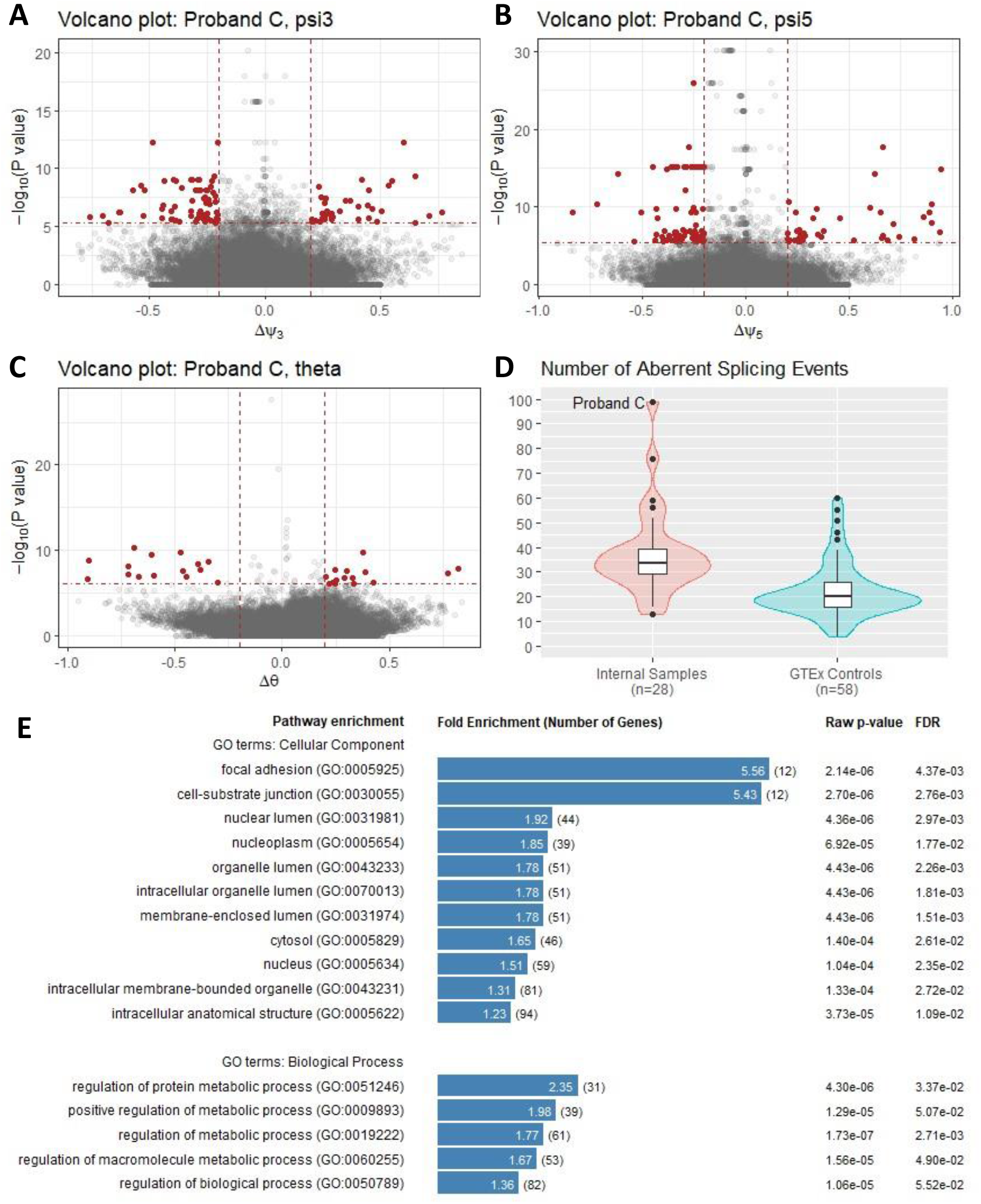
Whole transcriptome analysis of Proband C. (A-C) Volcano plot of aberrant splicing events detectd by three splicing metrics (psi3 for alternative splicing donors, psi5 for alternative splicing acceptors, and theta for aberrant splicing efficiencies) of FRASER in DROP. Red dots are significant aberrant splicing events with false discovery rate ≤ 0.05 (horizontal cut-off line) and magnititue ≥ 0.2 (vertical cut-off line). (D) Distrubution plot of the number of aberrant splicing event in indivduals of the internal RNA-seq samples (n=28) and GTEx controls (n=58). Proband C is a significant outlier comparing to the internal samples and external controls. (E) Pathway enrichment analysis by PANTHER. Three of the GO term for cell component are related to nucleus.

## Discussion

In this study, we present six individuals from five families with mild to severe global developmental delay, hypotonia, history of epilepsy or seizure, short stature, skeletal abnormalities and variable dysmorphic features, all carrying *de novo* and inherited alterations in *DDX39B*, a highly conserved RNA helicase belonging to the DExD/H-box RNA helicase family. All five variants were identified through trio ES (Supplemental information). In-depth *in silico* and *in vivo* analysis of the identified variants as well as blood transcriptomics profile of a patient, support the pathogenicity of the variants that are potentially contributing to an ultra-rare, novel neurodevelopmental syndrome.

DDX39B belongs the DExD/H-box RNA helicase family, a group of highly conserved proteins from yeast to humans. They contain a consensus amino acid sequence DExD or DExH in their Walker B motif (motif 2) within the helicase core domain, allowing for the ATP-dependent unwinding of RNA secondary structures.^20,23,26^ To date, multiple members of the DExD/H-box RNA helicase family, including *DDX3X* (MIM:300160), *DHX30* (MIM:616423), *DHX37* (MIM:617362), *DHX16* (MIM:602405), *DHX34* (MIM:615475), *DDX54* (MIM:611665), *DDX6* (MIM:618653) and *EIF4A2* (MIM:601102) have been casually linked to a heterogeneous group of neurodevelopmental disorders.^20,27–30^

DDX39B is an integral member of the TREX complex, a highly conserved protein complex that links transcription to mRNA export.^31–33^ Alterations in other members of the TREX complex (THOC2^8^ and THOC6^34^) have been linked to neurodevelopmental disorders, which include a spectrum of delayed development, moderate to severe intellectual disability, short stature, and dysmorphic facial features^8,29^. A review of the current literature on *THOC2*^35^ and *THOC6*^7,34,36–43^ related disease suggested several similarities (Table 1S). Short stature is reported in over 53% of all reported case descriptions, and at least 34% have abnormal cranium or forehead development in the form of a broad high forehead or microcephaly. Global developmental delay is a common feature that includes intellectual disability and speech delay (reported in 82% of *THOC2* and 100% of *THOC6*). Of interest, ventriculomegaly is found in Patient C and 4 of THOC2 and 4 of THOC6 reports, respectively. More comprehensive MRI assessment may yield common and consistent features in the three groups of diseases. Remarkably, lateral deviation at the metatarsophalangeal joint is a unique feature in our cohort and not seen in the *THOC2* and *THOC6* cohorts, comparing to overlapping or clinodactyly of toes reported in five patients with *THOC6* variants. In addition to its role in mRNA export, DDX39B, is also involved in pre-mRNA splicing by recruiting U2 snRNP at the pre-mRNA branchpoint.^44^ It is also possible that the variants described here also impede pre-mRNA splicing. More work is needed to delineate if *DDX39B* disease-causing variants result from defects in mRNA export, pre-mRNA splicing, or both.

The affected amino acid residues described here are highly conserved from yeast to humans. Residues Gly92 and Arg123 are also highly conserved in other DExD/H-box RNA helicases *DDX59* and *DDX6*, suggesting they are critical for proper protein function. Our *in silico* analysis suggests that the four missense *DDX39B* variants disrupt local protein folding and conformation. In the instance of p.(Gly92Asp), which occurs in the ATP binding pocket, we speculate that p.(Gly92Asp) results in impaired ATP hydrolysis. For p.(Arg123Gln), the loss of two polar bonds (Gly150 and Gln323) greatly impacts protein folding and possibly stability. While p.(Gly37Cys) and p.(Ser44Arg) fall outside characterized functional protein domains, we speculate that changing the wildtype residue’s size, biochemical properties and perturbing posttranslational modifications, in the case of p.(Ser44Cys), greatly perturbs protein function and structure.

Based on *in silico* analysis, the c.433-1G>T variant is expected to cause complete exon 5 skipping, resulting in a frameshift p.(Val145Thrfs11Ter) variant. The newly created termination codon occurs >600 nucleotides away from the last exon-exon junction; as such, we expect this allele to undergo nonsense-mediated mRNA decay (NMD) and is a null allele.^45–47^ Exon 5 encodes residues 145-206, which encodes most of the helicase ATP binding domain. If the mutant transcript escapes NMD and encodes a mutant protein, it would be missing >50% of the wildtype protein. Interestingly, Family 1, which segregates a canonical splice-site variant, has a milder phenotype than individuals with missense variants, although all are seemingly causing disease presumably through haploinsufficiency. This could be explained by leaky splicing or creating a new cryptic splice site. Visually scanning and using *in silico* predictions for possible cryptic splice sites failed to identify any potential cryptic splice sites. Given the variant impacts a canonical splice motif, it is not anticipated any wildtype splicing remains. An alternative and more attractive genotype-phenotype correlation could be the impact of a variant on the transcript and protein abundance.

*DDX39B* has a highly conserved paralog *DDX39A*. ^48,49^ *DDX39B* and *DDX39A* have been shown to have functional redundancy. ^48,50^ It has been shown that *DDX39B* needs to be tightly regulated as both overexpression and decreased expression is toxic in C. elegans^51^ and yeast.^52^ We speculate that null alleles lead to a less severe phenotype as *DDX39A* could partially substitute for *DDX39B*. Whereas, with missense variants where the *DDX39B* levels are not expected to change, the mutant protein is incorporated into the TREX complex and therefore compromises the entire function of the complex. Additional studies are needed to assess this hypothesis fully. *DDX39B* has a probability of loss-of-function intolerance (pLI) score of 1 based on gnomAD suggesting that the pathomechanism for *DDX39B* may be a haploinsufficiency and the loss of a single gene copy may cause the observed phenotypes. *DDX39B* also has a missense Z score of 4.564, suggesting that missense variants may not be tolerable. However, heterozygous copy number loss has been observed in multiple controls in the DGV database and large multigene genomic duplications and deletions have been reported in ClinVar. Together, these human population genetic data suggest that haploinsufficient or dominant-negative variants of DDX39B may create phenotypes. Both haploinsufficiency and dominant-negative mechanisms were initially proposed for *DDX6* (pLI=1, z=3.781), *DHX30* (pLI=1, z=5.296) and other recently described potentially haploinsufficient genes.^53^ However, a closer examination of human population genetic data for these genes reveals the presence of copy losses in normal controls. Therefore, we hypothesize that *DDX39B* variants, like the recently discovered DEAD/H box RNA helicases, are partial loss-of-function variants with a possible dominant-negative effect. Based on gene constraint metrics and the location of missense alterations described herein, dominant negative appears to be the most likely mechanism.

Previously, in *Drosophila* knock down of the *DDX39B* ortholog, *Dmel\Hel25E*, has been shown to inhibit growth and accumulate polyadenylated RNAs within the nucleus.^54^ We used *Drosophila* overexpression and rescue assays to better characterize three of the variants identified in this study, p.Gly37Cys, p.Gly92Asp, and p.Arg123Gln. In overexpression assays, wildtype DDX39B results in lethality, when ubiquitously expressed. In sharp contrast, the expression of the mutants do not cause lethality. In rescue experiments, neither reference nor variant can rescue fly loss of *Hel25E*, suggesting although highly conserved, *Hel25E and DDX39B* functions might not be completely redundant. Together, this data suggests that the p.Gly37Cys, p.Gly92Asp, and p.Arg123Gln alleles act as LoF. These data should be interpreted with caution given that human cDNA does not rescue, however, they do suggest some loss-of-function impact of the variants in this setting of *in vivo* overexpression.

Finally, the effect of *DDX39B* p.(Arg123Gln) variant is further supported by the disrupted transcriptomics profile identified in Proband C with RNA-seq. Blood transcriptome of the patient revealed 99 aberrant splicing events which is a significantly larger number than the internal samples and external GTEx controls (p<0.01). This transcriptional signature reflected a specific role of *DDX39B* in mRNA splicing regulation. We further evaluated the genes involved in the transcriptional signature. We found that most of the genes with aberrant splicing were protein encoding (89.9%), while the remaining affected genes were psudogenes and long non-coding RNA genes. An interaction enrichment among the protein-coding genes with aberrant splicing was identified (PPI enrichment p-value < 0.05) in the analysis with STRING database (https://string-db.org/, last accessed March 29, 2023), and DDX39B is located in the largest cluster in the derived STRING network. Enrichment analysis of gene ontology (GO) terms by the PANTHER Overrepresentation Test (Released 2022-10-13, http://pantherdb.org/webservices/go/overrep.jsp, last accessed April 2, 2023) revealed that the affected genes have an association with multiple cell components and metabolic processes (Figure 5E). The most enriched cellular component is focal adhesion and cell-substrate junction with fold change of 5.56 and 5.43 respectively. Another three enriched GO terms are related to the nucleus (namely nuclear lumen, nucleoplasm and nucleus) with fold changes ranged from 1.51 to 1.92, suggesting a potential disruption of essential cell components due to the identified *DDX39B* variant. The genetic conditions associated with the affected genes was further analyzed with the Clinical Genomic Database of the National Human Genome Research Institute (https://research.nhgri.nih.gov/CGD/, last accessed April 3, 2023). Among the 31 curated diseases caused by the affected genes, the most common manifestation categories are neurologic (n=21, 67.7%), followed by craniofacial (n=13, 41.9%) and musculoskeletal (n=13, 41.9%), which are potentially relevant to the phenotypes of our patient. The transcriptional signature can be further validated with future RNA samples from other patients.

In summary, the combined evidence of clinical features, *de novo* occurrences in the four most severe cases, evolutionary conservation of mutated residues, and in vivo functional effects strongly implicates *DDX39B* as the causative gene for the phenotype observed in our cohort. Moreover, the shared phenotypic characteristics among individuals with pathogenic variants in *DDX39B*, *THOC2*, and *THOC6* point towards a group of related disorders, which we propose to designate as ‘TREX-complex-related neurodevelopmental disorder’. Our findings also extend the current knowledge of the genetic contributions from DEAD-box RNA helicases and the TREX complex to neurodevelopmental syndromes. Further research is necessary to clarify DDX39B’s precise role during development, and more extensive cohort studies are required to explore the full range of clinical presentations and genotype-phenotype associations.

## Data and code availability

*DDX39B* variants reported herein have been deposited to ClinVar, temporary Submission ID: SCV003843896 for NM_004640.7:c.109G>T, SCV003843897 for NM_004640.7:c.275G>A, SCV003843898 for NM_004640.7:c.368G>A, SCV003843899 for NM_004640.7:c.132C>G, SCV003843900 for NM_004640.7:c.433-1G>T.

## Data Availability

DDX39B variants reported herein have been deposited to ClinVar, temporary Submission ID:
SCV003843896 for NM_004640.7:c.109G>T, SCV003843897 for NM_004640.7:c.275G>A, SCV003843898 for NM_004640.7:c.368G>A, SCV003843899 for NM_004640.7:c.132C>G, SCV003843900 for NM_004640.7:c.433-1G>T.

https://decipher.sanger.ac.uk

http://dgv.tcag.ca/

https://stuart.radboudumc.nl/metadome/

https://stuart.radboudumc.nl/metadome/

https://alphafold.com/

https://genematcher.org/

https://gnomad.broadinstitute.org/

https://matchmakerexchange.org/

https://www.omim.org/

## Acknowledgments

We thank all individuals, their families, and the referring physicians who submitted all clinical and genomic information. No additional compensation was received for these contributions. We extend our deep appreciation to the patient and his family for participating in this study. The authors thank the contributors to MyGene2, GeneMatcher, and other Matchmaker Exchange databases and the Genome Aggregation Database (gnomAD) and the groups that provided exome and genome variant data to these resources.

This study makes use of data generated by the DECIPHER community (a list of contributing centers is available from https://deciphergenomics.org/about/stats and via email from) and the DDD study. The DDD study presents independent research commissioned by the Health Innovation Challenge Fund [grant number HICF-1009-003], a parallel funding partnership between Wellcome and the Department of Health, and the Wellcome Sanger Institute [grant number WT098051]. The research team acknowledges the support of the National Institute for Health Research, through the Comprehensive Clinical Research Network. Those who carried out the original analysis and collection of the Data bear no responsibility for the further analysis or interpretation of the Data. This work was supported in part by the Indiana University Grand Challenge Precision Health Initiative, the Commissioned Paediatric Research at Hong Kong Children Hospital (PR-HKU-4) under the Health and Medical Research Fund of the Hong Kong Food and Health Bureau, and the Society for the Relief of Disabled Children.

## Web resources

CADD – Combined Annotation Dependent Depletion, https://cadd.gs.washington.edu/snv

DECIPHER, https://decipher.sanger.ac.uk

DGV – Database of Genomic Variants, http://dgv.tcag.ca/

MetaDome, https://stuart.radboudumc.nl/metadome/

AlphaFold, https://alphafold.com/

GeneMatcher, https://genematcher.org/

gnomAD, https://gnomad.broadinstitute.org/

Matchmaker Exchange, https://matchmakerexchange.org/

Online Mendelian Inheritance in Man, https://www.omim.org/

UCSC Genome Browser, https://genome.ucsc.edu/

https://research.nhgri.nih.gov/CGD/

